# Start Smart, Then Focus: Antimicrobial Stewardship Practice at One NHS Foundation Trust in England Before and During the COVID-19 Pandemic

**DOI:** 10.1101/2023.06.09.23291146

**Authors:** Rasha Abdelsalam Elshenawy, Nkiruka Umaru, Zoe Aslanpour

## Abstract

**Background:** Antimicrobial Resistance (AMR), a major global public health threat causing 1.2 million deaths, calls for immediate action. Antimicrobial stewardship (AMS) promotes judicious antibiotic use, but the COVID-19 pandemic increased AMR by 15%. Our study evaluated AMS implementation and inappropriate antibiotic prescribing before-the-pandemic (PD) and during-the-pandemic (DP).

**Methods:** This retrospective study examined medical records of adult patients (age 25 and above) admitted to an NHS Foundation Trust in England for respiratory tract infections (RTIs) or pneumonia in 2019 and 2020. Our objective was to evaluate antibiotic prescribing practices BP and DP in 2019 and 2020. Primary outcomes included evaluating the prevalence of inappropriate antibiotic prescribing and assessing the implementation of AMS using Public Health England’s ‘Start Smart, Then Focus’ (SSTF) toolkit. Reliable data extraction was ensured by two independent reviewers using a validated data extraction tool.

**Results:** A total of 640 patient records (320 from 2019 and 320 from 2020) were analysed. The mean age of enrolled adults was 74.3 years in 2019 and 76.2 years in 2020. COVID pneumonia showed a significantly higher odds ratio (OR) of 20.24 (95% CI 5.82 to 128.19, p-value<0.001). Inappropriate antibiotic prescribing, as per local guidelines, increased from 36% in 2019 to 64% in 2020 for the second course of antibiotics DP. Differences were observed in AMS interventions, with an OR of 3.36 (95% CI 1.30-9.25, p=0.015) for ‘Continue Antibiotics’ and an OR of 2.77 (95% CI 1.37-5.70, p=0.005) for ‘De-escalation’.

**Conclusion:** The COVID-19 pandemic significantly impacted antibiotic prescribing, increasing inappropriate use and posing risks of antimicrobial resistance. Factors influencing prescribing practices must be considered, and proactive measures, including updating the SSTF toolkit and developing an AMS roadmap, are needed to address the challenges of AMR in the context of evolving infectious diseases.

**KEY MESSAGES:** *WHAT IS ALREADY KNOWN ON THIS TOPIC:*

- ⇒ AMR is a major global health threat, called a silent pandemic, with the potential for 10 million annual deaths by 2050, equivalent to one death every three seconds.
- ⇒ Antimicrobial stewardship (AMS), promoting judicious antibiotic use, plays a pivotal role in combating AMR.
- ⇒ The COVID-19 pandemic led to a 15% rise in AMR and hospital-associated deaths during 2020.

*WHAT THIS STUDY ADDS:*

- ⇒ Evaluated the implementation of AMS before and during the COVID-19 pandemic in 2019 and 2020 across four seasonal time points.
- ⇒ Estimated the prevalence of inappropriate antibiotic prescribing in 2019 and 2020.
- ⇒ Identified factors influencing antibiotic prescribing upon admission and during the hospital stay.

*HOW THIS STUDY MIGHT AFFECT RESEARCH, PRACTICE AND/OR POLICY:*

- ⇒ Our study offered a comprehensive analysis of AMS implementation and identified the key factors that influence antibiotic prescribing and AMS application BP and DP. This critical understanding will be instrumental in shaping a strategic plan intended to improve antibiotic prescribing practices in acute care settings, thereby directing necessary updates and revisions in current policies.

## INTRODUCTION

The escalating prevalence of multi-drug-resistant infections worldwide presents an immense health threat, heightening morbidity, mortality, and economic consequences. The surge in multi-drug-resistant infections signifies a profound global health risk. In 2016, the O’Neill^1^ review highlighted an impending silent pandemic, foreseeing a staggering 10 million yearly deaths due to AMR by 2050, amounting to one death every three seconds. This alarming forecast highlights the urgent necessity for coordinated action, innovation, and collaborative efforts to prevent this impending public health crisis. In 2019, the World Health Organization (WHO) ^2^ categorised AMR among the top ten global public health threats necessitating immediate intervention. In 2019, the global death from AMR-related causes alarmingly increased to 1.2 million, emphasising the critical need for action.^3^

Antimicrobial stewardship, an organisational strategy advocating judicious antibiotic use, is pivotal to the UK’s Five-Year Antimicrobial Resistance Strategy.^4^ This initiative aims to enhance the quality and safety of patient care whilst significantly mitigating the emergence and spread of AMR. Public Health England (PHE)^5^ has acknowledged the essential role of AMS in combating AMR, providing the ‘Start Smart, Then Focus’ (SSTF) toolkit for AMS implementation in acute care settings. This approach promotes timely and responsible antibiotic use, prescribing effective antibiotics to treat infections, followed by an active ‘antibiotic review’ within 24-72 hours. The suggested SSTF method is applicable to all antibiotic prescriptions, thereby streamlining the process of antimicrobial prescribing. AMS encompasses strategies and interventions to improve antibiotic prescription appropriateness across all healthcare settings.^6 7^

The COVID-19 pandemic, triggered by the severe acute respiratory syndrome coronavirus-2 (SARS-CoV-2), originated in Wuhan, China, in December 2019 and swiftly spread worldwide.^8^ By June 2022, approximately 544 million people had tested positive for COVID-19, resulting in an estimated 6 million deaths.^9^ Recent research suggests that increased antimicrobial therapy during the pandemic may have contributed to the rise in resistant infections globally.^10^ In 2021, the Centres for Disease Control and Prevention (CDC) reported that the COVID-19 pandemic had resulted in a 15% increase in AMR and related deaths in hospitals in 2020, highlighting the need for further research and intervention to address this issue.^11^ Therefore, providing empirical data concerning the impact of the COVID-19 pandemic on antimicrobial prescribing and AMS is essential in reassessing and updating existing policies, along with the AMS roadmap. the AMS roadmap. This approach will, in turn, reduce the potential effects of future emergencies or crises on the AMS within acute care settings and will contribute to alleviating the threat of AMR. This cross-sectional retrospective study was conducted in an English NHS Trust in order to evaluate antibiotic prescribing and the impact of COVID-19 on AMS practices. Data were extracted from eight-time points across 2019 and 2020 before-the-Pandemic (BP) and During-the-Pandemic (DP).

## METHODS

### Study aims

1. To evaluate AMS implementation between BP and DP periods using the SSTF toolkit.
2. To determine the prevalence of inappropriately prescribed antibiotics BP and DP.
3. To identify factors influencing antibiotic prescribing and AMS implementation in both BP and DP.

### Study design

A retrospective cross-sectional analysis was undertaken to estimate the prevalence of inappropriate antibiotic prescribing in adult patients aged 25 years and above who were admitted to one NHS Foundation Trust in England. This secondary care provider serves approximately 400,000 people and consists of about 742 beds. A comprehensive literature review was undertaken to determine the most suitable tool for the investigation.

### Sample size

As indicated by PHE, it was estimated that 20% of all antibiotics prescribed in the UK may be inappropriate.^12^ The sample size for this study was determined, taking into consideration both this percentage and relevant literature. The statistical software, Minitab, was employed to calculate the sample size, incorporating variables such as the size of the population, a margin of error set at 10%, and a confidence interval of 95%. The data were drawn from medical records at eight different time points to allow for the variation in antibiotic prescribing across seasons. This included four baseline points BP and four points DP. Data were extracted from 320 patient records from 2019 (BP) and 320 from 2020 (DP), totalling 640 records. Four-time points were employed each year, using a systematic sampling methodology, allowing for a sample of 80 patients at each point.

### Study population (inclusion/exclusion criteria)

A stratified sampling strategy was employed to ensure maximum diversity among the included Medical Records (MRs). The inclusion criteria comprise the following: (i) adult patients aged 25 years and older; (ii) pregnant women and immunocompromised patients; (iii) patients admitted to the Trust; (iv) patients admitted in 2019 and 2020; and (v) patients prescribed antibiotics for upper respiratory tract infections (URTIs) or pneumonia. However, patients who spent less than 48-72 hours in the Accident and Emergency (A&E) department, patients who were not prescribed antibiotics, and children were excluded from this study.

### Data source

The main author (RAE) extracted the data from the patient’s electronic medical records within the Trust, adhering to the study’s inclusion and exclusion criteria.

### Data collection

Data was collected from the patient’s electronic medical records within the Trust in accordance with the study’s inclusion and exclusion criteria. The data collection process for each patient’s medical record took about 45 minutes. Data was gathered from eight-time points, with four-time points BP: (i) March (Spring 2019); (ii) June (Summer 2019); (iii) September (Autumn 2019); and (iv) December (Winter 2019). Additionally, four-time points occurred DP: (i) March (Spring 2020) - the first wave of COVID-19; (ii) June (Summer 2020) - the first lockdown; (iii) September (Autumn 2020) - the second wave of COVID-19; and (iv) December (Winter 2020) - the vaccination rollout.

### Data Extraction

A data extraction tool was employed to obtain the necessary data from patients’ medical records. A Mind Map was created to aid in organising the data extraction tool in relation to the antibiotic use process and the PHE toolkit for AMS (online supplemental material 1). In order to extract data from patients fitting the inclusion criteria, access to the Trust’s electronic system was required. Prior to commencing ‘Data Extraction’, the main author completed training modules for all these systems and subsequently gained access to them. The data extraction tool was prepared in order to obtain the necessary information from the patient’s medical records. The AMS Data Extraction Tool was prepared, encompassing demographic information, primary diagnosis, SSTF criteria, AMS interventions, investigations, and patient outcomes. The extraction process took approximately 45 minutes per patient medical record from the main author to gather the required data (online supplemental material 2).

### Pilot study

The research student conducted a pilot study, extracting data from 10 medical records for each time point, totalling 80 patient medical records in 2019 and 2020. The pilot study aimed to offer an initial description of the data and evaluate the feasibility of the data extraction tool in addressing the research questions. Both descriptive and statistical data were expected to be included. The pilot study’s results indicated that the data extraction form adequately addressed all the study objectives. However, due to the small sample size of the pilot study, not all statistical analyses could be applied. Performing statistical tests for relationships (associations and correlations) was not possible. Additional data was necessary to calculate the prevalence of antibiotic prescribing and AMS implementation. The study analysis did not incorporate data generated and extracted from the pilot test.

### Validity and Reliability of the Data Extraction Tool

The primary author (RAE) utilised literature and the AMS PHE Toolkit, in order to construct the data extraction tool. The elements within the tool were recognised and consented to through the authors’ discussion. To validate the data extraction tool, RAE and an AMS pharmacist at the research site each separately extracted data from 1% of the sample (four patient records). An agreement rate of 80% or higher served as a measure of the tool’s validity. Furthermore, to assess the tool’s reliability, both RAE and the AMS pharmacist independently extracted data from a similar 1% of the sample (four records). Inter-rater reliability was determined by examining the percentage of agreement in the data extracted independently. Any disagreements were resolved through dialogue.

### Data Analysis

In this study, the prevalence of inappropriate antibiotic prescribing was evaluated based on the local antimicrobial prescribing guidelines. This was done by identifying the initial antibiotic selected for each patient according to the main diagnosis of RTIs or pneumonia and comparing it with the hospital antimicrobial prescribing guidelines to compute the percentage of inappropriate prescriptions before and during the pandemic. Additionally, AMS implementation was measured using the PHE AMS Toolkit.^5^ Demographics and clinical attributes were described using descriptive statistics, and AMS intervention prevalence was also determined. Advanced statistical analysis was conducted using IBM SPSS Statistics version 22.0,^13^ RStudio version 2022, and R version 4.2.2.^14^ A framework for data analysis was established for data analysis (online supplemental material 3).

### Patient characteristics

The number of patients, mean age, gender, admission speciality, patient classification, and discharge method. Further data was collected on the percentage of inappropriate antibiotic prescribing before and during the COVID-19 pandemic. Additionally, the number of patients admitted with RTIs as their main diagnosis according to each chapter of the ICD-10 was recorded. All these data were connected to the measure of AMS interventions. The average characteristics of the patients were determined by considering the length of their hospital stays (LOS).

### Data anonymisation and access

In this retrospective research, patient-identifiable data was accessed without explicit consent. Post HRA approval, the corresponding author communicated with the AMS pharmacist within Trust to initiate the study. The AMS pharmacist liaised with the coding team to prepare a list of RTI diagnoses using the ICD-10 system, corresponding to the study’s timeline. Ensuring adherence to the National Opt-out Act, they also interacted with the Information Governance Team. Post-data extraction, the collaborator anonymised the dataset before handing it to the author. Anonymised data collection and processing was fair and lawful in line with General Data Protection Regulation (GDPR) principles, Caldicott Guardian, and Trust protocols.

### Patient and public involvement

The study protocol was sent to representatives of the Citizens Senate, a patient care organisation with a good representation of many older people. They reviewed it and provided feedback.

### Registration

This study has been registered in the ISRCTN registry, which is a primary registry recognised by WHO and ICMJE that accepts all clinical research studies.^15^

## RESULTS

### Clinical and Demographic Characteristics

A retrospective investigation was conducted on 640 medical records of patients admitted to the Trust. The demographics of these patients are detailed in Table 1. None of the variables in the comprehensive analysis yielded statistical significance between 2019 and 2020. The age span of patients admitted with RTIs in 2019 and 2020 ranged from 25 to 99 years, with a marginal variation in gender distribution; in 2019, 160 (49%) were female, slightly increasing to 159 (50%) in 2020. Similarly, the mortality rates documented in Patient Outcomes consistently remained at 15% throughout the two-year period. The LOS represented 13% and 12%, respectively, over these two years. Remarkably, the average LOS was nearly congruent between the two years. In 2019, the LOS extended from 1 to 119 days, whereas in 2020, it was between 1 to 97 days. The standard deviation stood at 16 in 2019 and slightly reduced to 13 in 2020 (Table 1).

**Table 1.**
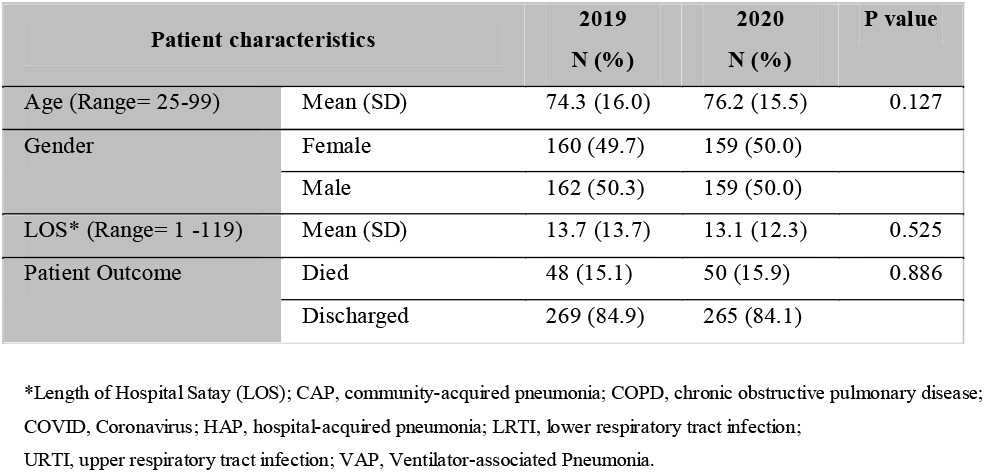
Characteristics of the patient demographics admitted before the COVID-19 pandemic (n=320) and during the pandemic (n=320) (in 2019 and 2020).

| Patient characteristics |  | 2019<br>N (%) | 2020<br>N (%) | P value |
| --- | --- | --- | --- | --- |
| Age (Range= 25-99) | Mean (SD) | 74.3 (16.0) | 76.2 (15.5) | 0.127 |
| Gender | Female | 160 (49.7) | 159 (50.0) |  |
|  | Male | 162 (50.3) | 159 (50.0) |  |
| LOS* (Range= 1 -119) | Mean (SD) | 13.7 (13.7) | 13.1 (12.3) | 0.525 |
| Patient Outcome | Died | 48 (15.1) | 50 (15.9) | 0.886 |
|  | Discharged | 269 (84.9) | 265 (84.1) |  |

\*Length of Hospital Stay (LOS); CAP, community-acquired pneumonia; COPD, chronic obstructive pulmonary disease; COVID, Coronavirus; HAP, hospital-acquired pneumonia; LRTI, lower respiratory tract infection; URTI, upper respiratory tract infection; VAP, Ventilator-associated Pneumonia.

### Antimicrobial Stewardship: Start Smart - Then Focus

The term “Start Smart” denotes the initial stage of antibiotic administration.^5^ The discrepancy in the appropriateness of antibiotic prescriptions before and amidst the COVID-19 pandemic seems statistically insignificant. Notably, while the age category overall was not statistically significant, the age category of 75-84 encompassed the greatest number of admissions in 2019 and 2020 (with 521 and 389 admissions, respectively). Age and gender do not appear to impact antibiotic prescribing patterns significantly. Similarly, there is no significant change in the documentation of antibiotic allergies before and during the pandemic. However, a notable difference was observed in the occurrence of side effects, with an OR of 7.23 (95% CI 1.54 to 53.37, p-value=0.023).

Additionally, there were several factors that influenced this initial antibiotic prescribing ‘Start Smart’, including the main diagnosis. For example, Community-Acquired Pneumonia (CAP) was the predominant diagnosis in approximately 128 pre-pandemic patients. Still, the difference in the number of CAP patients pre- and post-pandemic is statistically insignificant. Interestingly, within the data procured from the study population, the severity risk assessment, CURB-65 score for CAP, was only reported in three patient records.^16^ The presence of unclear diagnoses, such as Upper Respiratory Tract in Infections (URTI), Lower Respiratory Tract Infections (LRTI), and Pneumonia, influence the appropriate choice of antibiotics at the time of admission. Notably, COVID pneumonia demonstrates a statistically significant difference between the years 2019 and 2020, with an odds ratio (OR) of 20.24 (95% CI 5.82 to 128.19, p-value<0.001). With respect to comorbid conditions, no notable differences were detected between the periods before and during the pandemic. However, Hypercholesterolemia showed a significant difference with an OR of 1.90 (95% CI 1.14 to 3.20, p-value=0.014). Heart failure demonstrated an OR of 2.06 (95% CI 1.23 to 3.52, p-value=0.007), kidney diseases exhibited an OR of 0.52 (95% CI 0.32 to 0.84, p-value=0.008), and asthma showed an OR of 0.50 (95% CI 0.25 to 0.95, p-value=0.038). Moreover, no significant differences were discernible in terms of antibiotic therapy duration, whether it was short-term (less than or equal to 3 days) or long-term (greater than or equal to 6 days), both BP and DP. In accordance with local guidelines, the prevalence of inappropriate antibiotic prescribing was identified as 50% in 2019 before the pandemic and maintained a similar rate during the pandemic in 2020, at 49%. (Table 2).

**Table 2.**
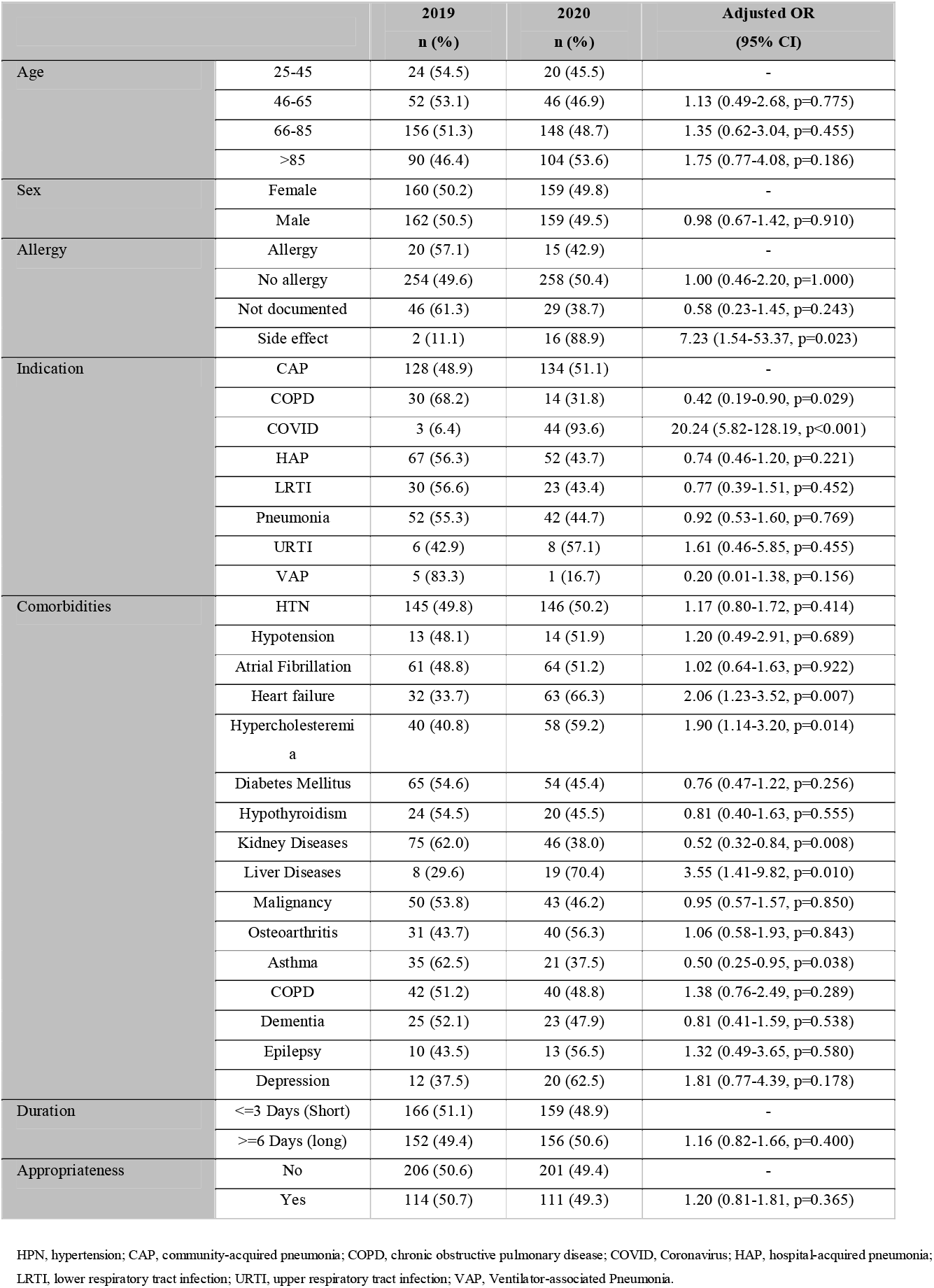
Adjusted ORs indicate the change in the odds of factors affecting the ‘Start Smart’ initial antibiotic prescribing in patients admitted with RTIs before and during the COVID-19 pandemic (2019 and 2020).

Table 3 provides an overview of factors impacting the ‘Then Focus’ antibiotic prescribing in patients with RTIs before and during the COVID-19 pandemic.^5^ No significant differences were observed in laboratory tests like White Blood Cells (WBCs), C-reactive protein (CRP), and Serum Creatinine. However, Chest X-ray radiological examinations presented notable results in diagnosing pneumonia before and during the pandemic, with an OR of 1.75 (95% CI 1.04 to 2.97, p-value=0.037). The Days of Antibiotic Review over this period didn’t show any significant changes. Concerning the AMS interventions, a meaningful difference was observed in the ‘Continue Antibiotics’ and ‘De-escalation’ decisions, with an OR of 3.36, 95% CI 1.30-9.25, p=0.015, and an OR of 2.77, 95% CI 1.37-5.70, p=0.005, respectively. In accordance with local guidelines, the percentage of inappropriate antibiotic prescriptions rose from 36% in 2019 (BP) to 64% in 2020 (DP).

**Table 3.**
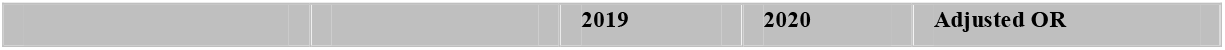
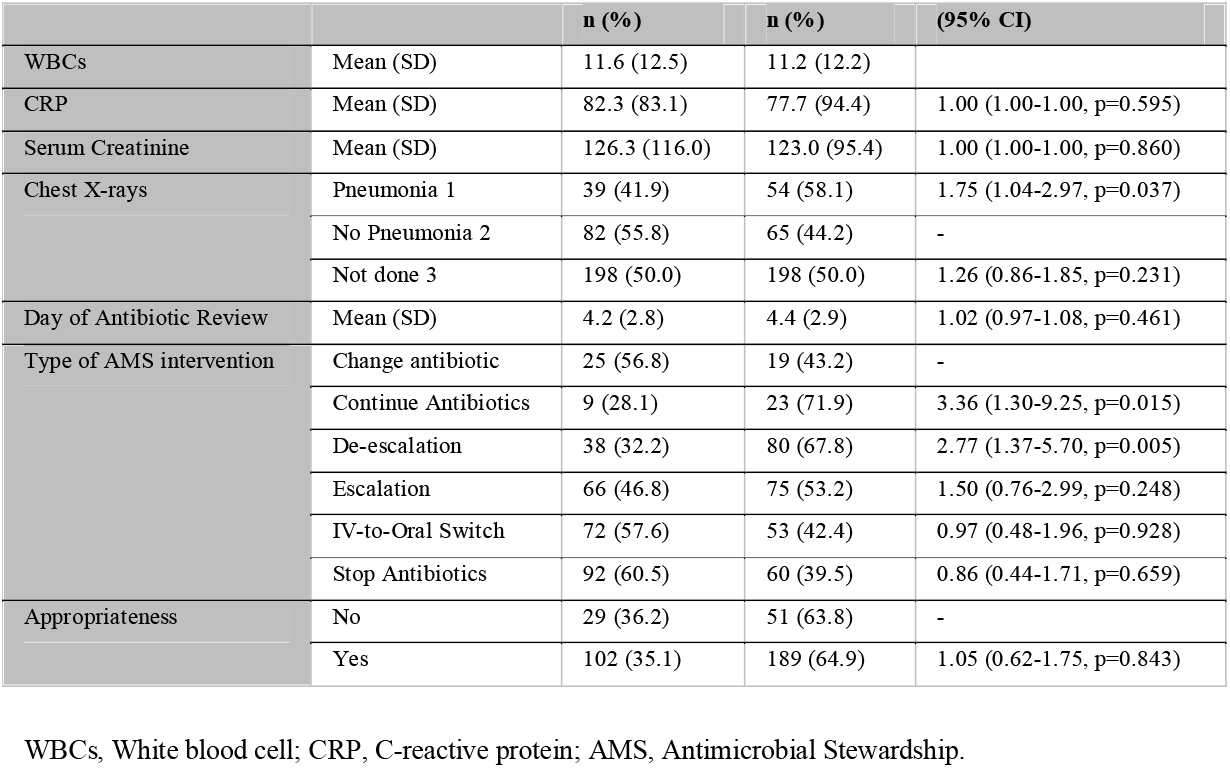
Adjusted ORs indicate the change in the odds of factors affecting the ‘Then Focus’ criteria of antibiotic prescribing in patients with RTIs before and during the COVID-19 pandemic (2019 and 2020).

|  |  | n (%) | n (%) | (95% CI) |
| --- | --- | --- | --- | --- |
| WBCs | Mean (SD) | 11.6 (12.5) | 11.2 (12.2) |  |
| CRP | Mean (SD) | 82.3 (83.1) | 77.7 (94.4) | 1.00 (1.00-1.00, p=0.595) |
| Serum Creatinine | Mean (SD) | 126.3 (116.0) | 123.0 (95.4) | 1.00 (1.00-1.00, p=0.860) |
| Chest X-rays | Pneumonia 1 | 39 (41.9) | 54 (58.1) | 1.75 (1.04-2.97, p=0.037) |
|  | No Pneumonia 2 | 82 (55.8) | 65 (44.2) | - |
|  | Not done 3 | 198 (50.0) | 198 (50.0) | 1.26 (0.86-1.85, p=0.231) |
| Day of Antibiotic Review | Mean (SD) | 4.2 (2.8) | 4.4 (2.9) | 1.02 (0.97-1.08, p=0.461) |
| Type of AMS intervention | Change antibiotic | 25 (56.8) | 19 (43.2) | - |
|  | Continue Antibiotics | 9 (28.1) | 23 (71.9) | 3.36 (1.30-9.25, p=0.015) |
|  | De-escalation | 38 (32.2) | 80 (67.8) | 2.77 (1.37-5.70, p=0.005) |
|  | Escalation | 66 (46.8) | 75 (53.2) | 1.50 (0.76-2.99, p=0.248) |
|  | IV-to-Oral Switch | 72 (57.6) | 53 (42.4) | 0.97 (0.48-1.96, p=0.928) |
|  | Stop Antibiotics | 92 (60.5) | 60 (39.5) | 0.86 (0.44-1.71, p=0.659) |
| Appropriateness | No | 29 (36.2) | 51 (63.8) | - |
|  | Yes | 102 (35.1) | 189 (64.9) | 1.05 (0.62-1.75, p=0.843) |

WBCs, White blood cell; CRP, C-reactive protein; AMS, Antimicrobial Stewardship.

This research further highlights the decisions related to antibiotic prescribing and AMS interventions implemented by The Start Smart – Then Focus AMS toolkit.^5-7^ This analysis examines how these decisions were modified in light of seasonal changes, as well as throughout the COVID-19 pandemic in the years 2019 and 2020. The present study observed a significant difference among AMS interventions in all seasons, as demonstrated by Figure 1, which depicts a bar chart for the AMS interventions in 2019 and 2020. Specifically, the De-escalation AMS intervention showed an increasing trend at the start of the COVID pandemic, with 14 out of 80 patients in Dec-2019, followed by a slight increase to 20 out of 80 patients in March 2020 and 25 out of 80 patients in Dec-2020. Meanwhile, the ‘Escalation’ AMS intervention showed the most significant rise in Sep-2020. Conversely, the Stop Antibiotics intervention experienced a decline in usage during 2020, being equally applied in March 2020, June 2020, and December 2020 (16 of 80 patients). The study found that during the pandemic’s onset in December 2019, the Stop Antibiotic and IV-to-Oral switch interventions were the most applied, administered to 17 and 25 of 80 patients, respectively. In contrast, in December 2020, the De-escalation intervention became the second most frequently used intervention. However, in March 2020, during the first wave of the pandemic, there was a contradiction in the AMS implementation, with both Escalation and De-escalation interventions being the most commonly used interventions (Figure 1).

**Figure 1.**
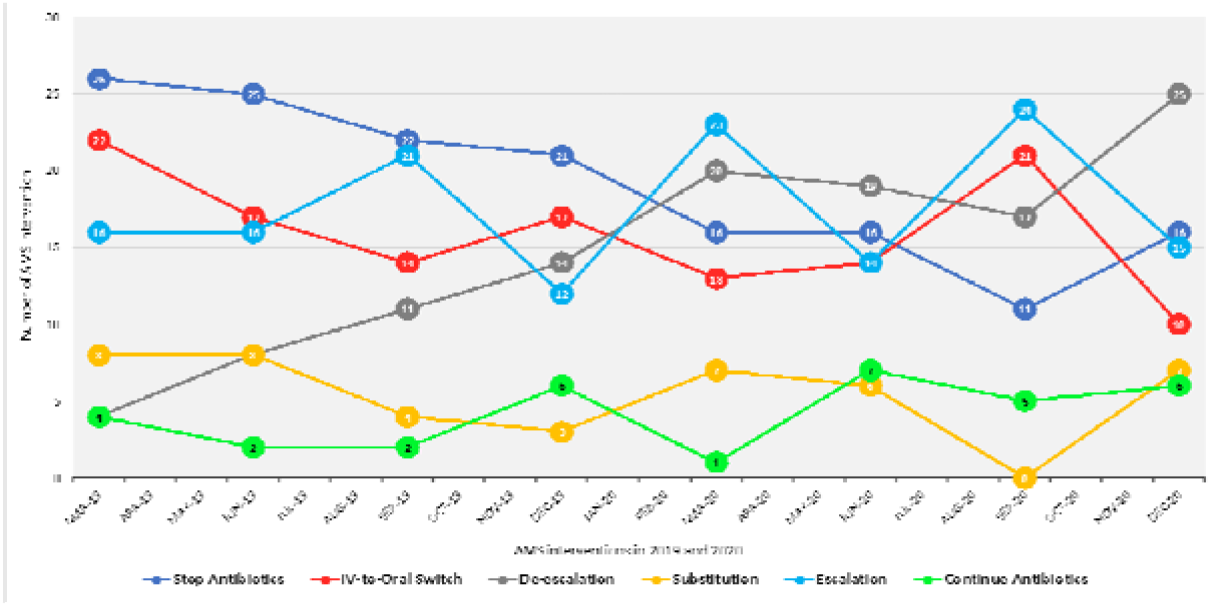
Antimicrobial Stewardship Interventions Before and During the COVID-19 Pandemic (2019 - 2020).

## DISCUSSION

### Principal findings

The study found multiple factors influencing initial antibiotic prescription at admission (‘Start Smart’), such as patient age, allergies, main diagnosis, and co-morbidities. According to local guidelines, a clear diagnosis was key for initial antibiotic selection. In addition, the use of severity risk assessment is important to identify the severity of infection, such as the CURB-65 score with CAP is important to select the right antibiotic was only found in three patient cases. This data scarcity may influence antibiotic prescribing appropriateness for patients diagnosed with CAP.^16^ About 50% of prescriptions were deemed inappropriate BP and DP. This rose to 63% for the second antibiotic course (‘Then Focus’) during the pandemic. Lab tests, including WBCs, PCR, Serum Creatinine, and chest X-rays, were significant in prescribing decisions. Antimicrobial Stewardship Interventions shifted during COVID-19. ‘Stop Antibiotics’ and ‘IVOS’ usage dropped with the first lockdown, while ‘Escalation’ increased. Upon the release of the NICE guidelines on Pneumonia, ‘Escalation’ decreased.^17^ There’s a pressing need for better awareness and education about Antimicrobial Stewardship interventions, emphasising the role of ‘IVOS’, ‘De-escalation’, and ‘Stop’ inappropriate antibiotic use relative to patient conditions.

### Comparison with other studies and clinical implications

This retrospective analysis evaluated the admissions of patients with RTIs during both BP and DP. It also highlighted the inplementation of AMS as a crucial part of the UK’s Five-Year AMR strategy in order to enhance patient care and combat AMR.^19^ It involved improving antibiotic prescribing, using a SSTF approach for antimicrobial stewardship.^5^ As previously stated, while the toolkit proved invaluable for analysing AMS in this study, it necessitated additional revisions to encompass other variables that influence the prescription of antibiotics, specifically the initial course upon admission, known as “Start Smart” and subsequent course(s) after hospitalisation, referred to as “Then Focus.” By incorporating these updates, we aim to ensure the long-term viability of implementing AMS, particularly in emergency situations, while concurrently reducing AMR.

A 2021 study in England found that antibiotic prescribing patterns changed during the COVID-19 pandemic, with more early prescriptions. Different infection types were affected differently, and AMS was compromised. Future adaptations in infection management and stewardship are necessary.^18^ The COVID-19 pandemic significantly influenced the number of patients admitted with RTIs. An increase in admissions in December 2019 could be attributed to the rapid spread of COVID-19 and its impact on respiratory health. A subsequent decline in March 2020 and June 2020 coincided with public health measures and the second national lockdown.^19^ In a 2023 study conducted in England, high rates of antibiotic prescribing were observed alongside low rates of confirmed respiratory infections through cultures. Nearly one-third of patients received multiple antibiotic courses, and highlighting also the impact of COVID-19 on antimicrobial stewardship.^20^ Interestingly, another separate 2019 UK study discovered that antibiotic prescribing often deviated from guidelines, particularly for URTIs. Future interventions should focus on optimising the rational use of antibiotics. ^21^ Our study indicated that clear diagnoses, such as URTIs, LRTIs, and pneumonia, influenced the appropriate selection of antibiotics upon admission.

In a 2021 study in Manchester, no significant differences were observed between shorter and longer antibiotic courses in infection-related hospitalisations, indicating their equal effectiveness. ^22^ Similarly, our study found no notable disparities in antibiotic therapy duration, whether short-term (≤3 days) or long-term (≥6 days), both before and after the pandemic. In a 2020 UK study, elevated CRP levels were found to predict bacterial and viral pathogens independently. However, their value, in addition to sputum purulence, was inconclusive.^23^ Our study emphasised the importance of additional investigations like WBCs and Chest X-rays to identify pneumonia patients who may benefit from antibiotics and ensure appropriate prescribing practices.

The COVID-19 pandemic has emphasised uncontrolled infectious diseases’ economic and societal impact, resembling predictions about AMR. Understanding the effects of changed antibiotic use, health-seeking behaviour, and infection control on AMR is crucial to promote good practices and prioritising research.^24^ Implementing stewardship programs should prioritise core strategies and focus on their effectiveness before incorporating supplemental approaches. Quality indicators and improvement projects help maintain the sustainability of antimicrobial stewardship implementation, especially during the pandemic.^6^ While healthcare professionals are dealing with the challenges of COVID-19, the ongoing crisis of AMR should not be neglected. Addressing AMR proactively can prevent future reactivity, similar to our response to COVID-19. ^25^ In a 2022 Lancet study, the antibiotic review kit intervention reduced antibiotic use among adult acute general medical inpatients. The COVID-19 pandemic likely influenced the inconsistent effects on mortality.^26^ Hospitals should adopt the antibiotic review kit to curb antibiotic overuse. Although there was no significant difference in the day of antibiotic review pre and during the pandemic, most reviews occurred after four days in both periods. This highlights the significance of multidisciplinary team ward rounds by AMS team for proper antibiotic review and decision-making for patients. SSTF audits yielded significant outcomes and valuable insights for other hospitals, resulting in improved antibiotic prescribing practices. Furthermore, incorporating other quality improvement methods can effectively enhance antibiotic prescribing in a sustainable manner. ^27-28^

### Strengths and limitations

Our study’s main strength lies in providing a comprehensive understanding of AMS both before and during the COVID-19 pandemic. This study contributes empirical data to evaluate antibiotic prescribing practices in relation to seasonal variations and the impact of the pandemic on antibiotic prescribing at one English NHS Trust in Great Britain. The study’s robust methodology, rigorous follow-up, and utilisation of a well-established retrospective review process, supported by a validated data extraction tool, ensure a thorough quantitative examination of AMS before and during the COVID-19 pandemic, thus minimising the likelihood of overlooking significant aspects in studying AMS implementation.

Our study does exhibit several limitations. Firstly, the study was conducted in an acute care setting within a secondary care hospital, limiting the generalisability of the findings to primary care settings. Furthermore, the study focused primarily on adult patients admitted during the years 2019 and 2020, predominantly within the age range of 75-84. We excluded patients under the age of 25 and children from this study. Additionally, among the enrolled patients, we specifically considered those with RTIs, thereby restricting the applicability of our findings to other types of infections. Finally, despite the large sample size, our study only examined the first and second courses of antibiotics. Notably, there was a reduction in the maximum LOS from 119 days in 2019 to 97 days in 2020. However, this study did not include other courses of antibiotics in patients with prolonged LOS, necessitating further investigation. However, our study offers valuable insights into the implementation of AMS. It is important to acknowledge and consider these limitations. Additional research should be conducted to address the identified gaps and broaden the scope of exploring healthcare professionals’ knowledge, perception and attitudes towards antibiotic prescribing during the COVID-19 pandemic in acute care settings.

## Conclusion

The COVID-19 pandemic has significantly impacted antibiotic prescribing patterns, increasing the risks associated with AMR and patient outcomes. Therefore, consistently implementing AMS measures is crucial to ensure appropriate antibiotic use and mitigate AMR. Factors influencing antibiotic prescribing upon admission, including patient age, allergies, main diagnosis, co-morbidities, and risk assessment, must be considered. In addition to the other factors of clinical assessment with the prolonged patient stay in the hospital, such as Lab tests, including WBCs, PCR, Serum Creatinine, and chest X-rays, has to be considered. Long-term effectiveness of antimicrobial stewardship requires continuous updates to the SSTF toolkit, particularly for emergency or crisis situations, to effectively combat AMR. Proactive measures, such as the development of tools or roadmaps, are necessary to facilitate antimicrobial stewardship interventions in acute care settings and address the challenges posed by AMR, especially in the context of evolving infectious diseases like COVID-19.

## Data Availability

The individual patient records cannot be shared by the investigators under the data use agreement with the NHS Trust studied in this paper.

## Contributors

RAE was responsible for data acquisition, with the study design and conceptualisation developed collaboratively by RAE, NU, and ZA. RAE carried out the literature review under NU and ZA’s supervision and further extracted relevant electronic data from patient records. The project dataset was constructed by RAE, who also verified, accessed, and analysed the data, guided significantly by NU and ZA. RAE produced the initial draft of the study, guided by NU and ZA. All authors contributed to data interpretation and the preparation and revision of the manuscript, sharing equal responsibility for the final decision to submit and approval of the final manuscript. ZA acted as the guarantor for the overall content of the research. Data were anonymised before analysis and securely stored within the University of Hertfordshire’s (UH) dual secure system. RAE analysed the anonymised data using UH’s dual secure system.

## Competing interests

All authors (RAE NU and ZA) have no competing interests.

## Patient consent for publication

Not applicable.

## Ethics approval

Ethical approval for this study was granted by the Health Research Authority (HRA), with the Research Ethics Committee (REC) assigning reference number 22/EM/0161. In compliance with this approval, the study protocol underwent review and received approval from the University of Hertfordshire (UH) ethics committee under the reference LMS/PGR/NHS/02975.

## Provenance and peer review

Commissioned; internally and externally peer-reviewed.

## Data availability statement

Data is not publicly available. The data, derived from patient records, is under a data usage agreement with the National Health Service Trust studied in this paper and cannot be shared by the investigators.

## Notes

### Competing Interest Statement

The authors have declared no competing interest.

### Clinical Protocols

https://www.isrctn.com/ISRCTN14825813

### Funding Statement

This research study has no fund

### Author Declarations

East Midlands Leicester South Research Ethics Committee (REC) gave ethical approval for this work - REC Reference: 22/EM/0161 University of Hertfordshire Ethics Committee gave ethical approval for this work, under the reference LMS/PGR/NHS/02975.

