## Supplement materials 1-3 for "Start Smart, Then Focus: Antimicrobial Stewardship Practice at One NHS Foundation Trust in England Before and During the COVID-19 Pandemic"

**‌** **Supplemental material 1: Mind Map organises data extraction for Antimicrobial Stewardship (AMS).**

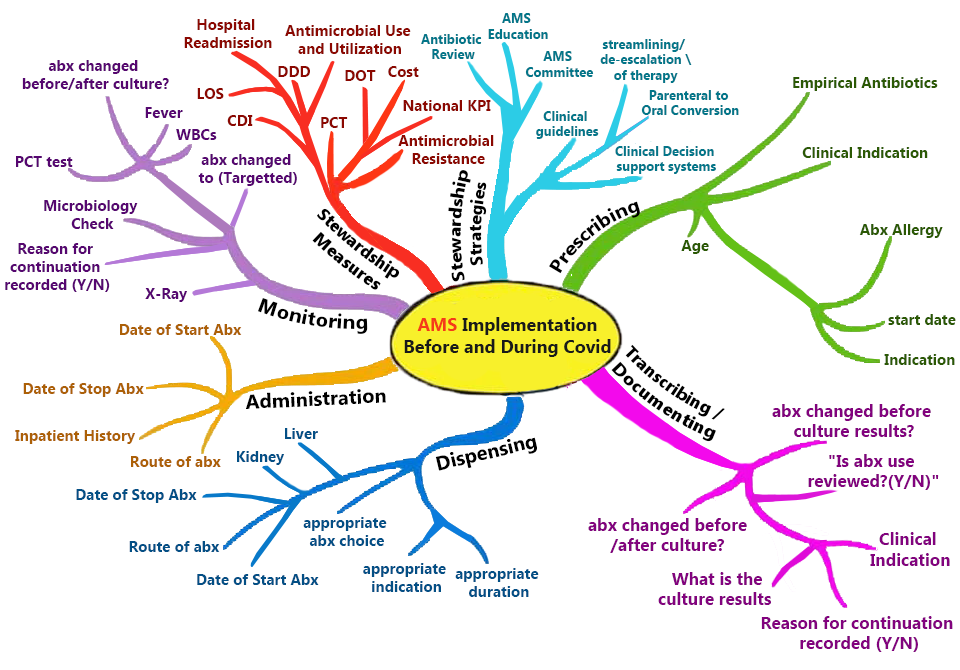

**Supplemental material 2: Data Extraction Tool**

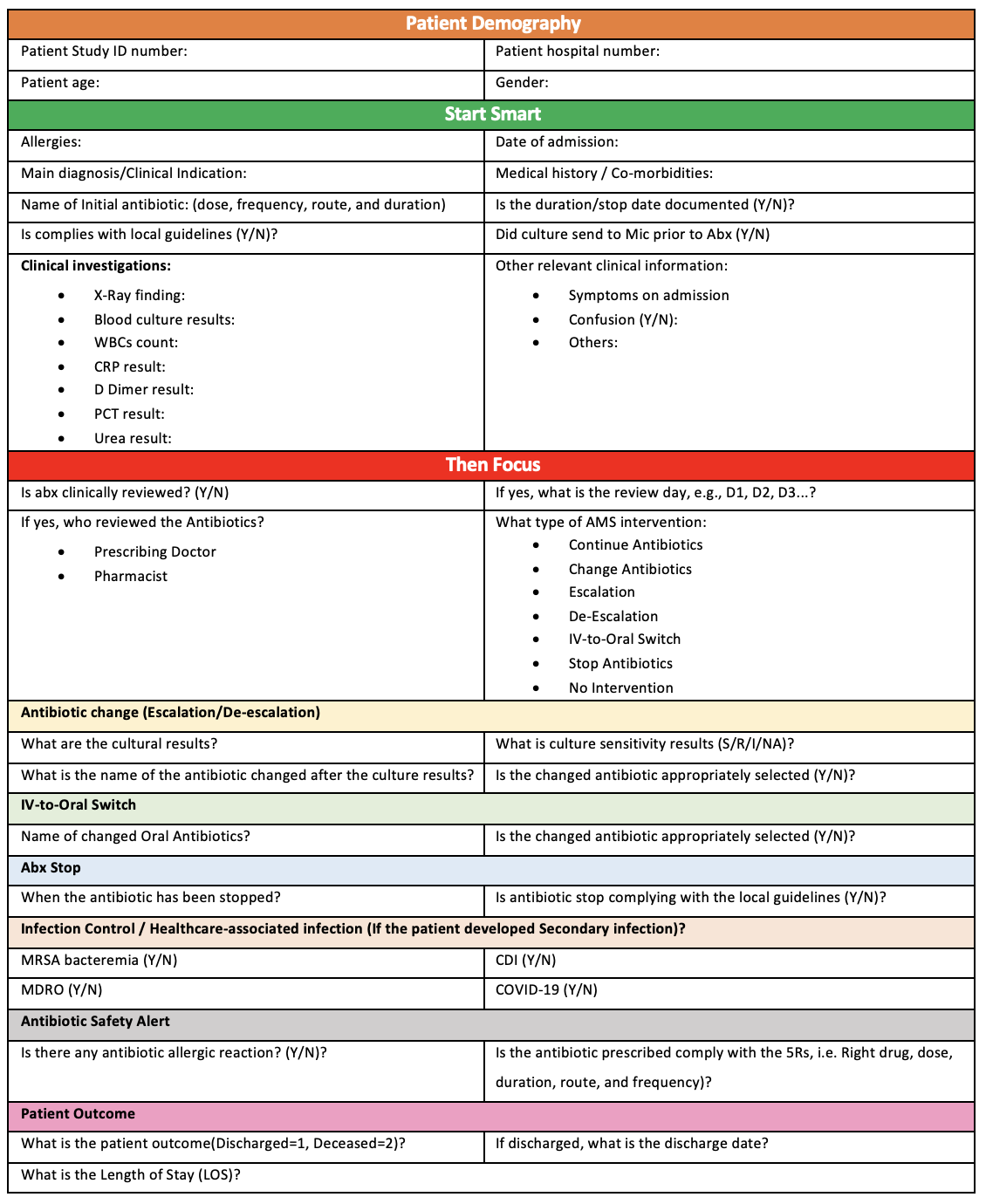

**Supplemental material 3: Framework established for data analysis.**

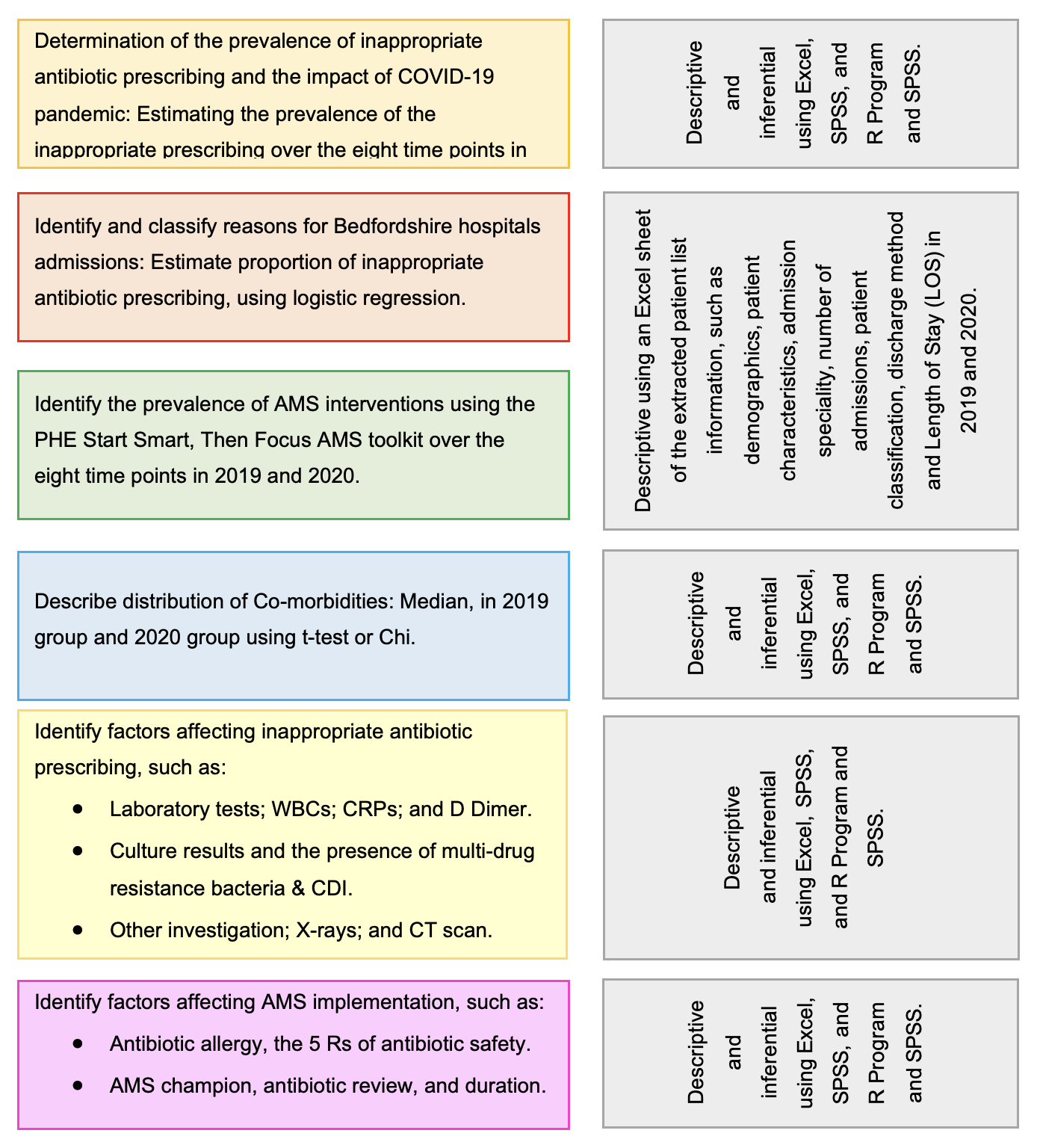

‌

‌
